# Locating heterogeneity in Menstrual Hygiene Scheme impact on Indian adolescent girls

**DOI:** 10.1101/2023.09.21.23295882

**Authors:** Olivia Sarkar, Arijita Dutta

**Affiliations:** Department of Economics, University of Calcutta, India

## Abstract

India has seen Menstrual Hygiene Scheme for 15-19 aged adolescents as a mass movement in 2011 with Intervention, Education, Communication (IEC) interventions and subsidized menstrual product. While the phenomenal increase in hygienic menstrual usage during 2010-20 is celebrated, it leaves us to question if this is the MHS in play or a gradual transition due to overall developmental drive. As the age-cohorts move out of the beneficiary net over time, sustainability of the program can only be reflected if there is a behavioral change which is captured through difference in choice patterns over duration of program exposure. Using triple difference-in- difference methodology on unit level data from NFHS 4, the results find that overall the program fails to mark any significant effect on choice of hygienic product in India. However, the low rung of the cohorts in terms of education and media exposure reap the benefit of the scheme, leaving the better-off counterparts unaffected. This concludes that the issues targeted through the scheme is holistic to reach the lowest strata of the society, but a proper choice of target cohort can considerably reduce the project cost, free it of supply side inefficiencies and enhance targeting of IEC techniques, thereby making the scheme more effective.

## Introduction

Recently, the discourse on menstrual health has been the top of talks for its contribution towards multi-dimensional inequality in a women’s life [1–5]. From driving absenteeism in workplace, dropouts from school to morbidities associated with menstruation, it has augmented the persistent gender inequality. To worsen the scenario of menstrual experience, the associated taboos and shame invariably across religions, menstruation cycle acts as a hindrance to attaining sustenance, empowerment and health equality [6–9]. Restriction to access of food, shelter and religious rights in family houses are what a woman faces monthly as part of their menstrual experience. The periodic shaming and associating taboos categorizing women as impure, belittles. Thereby, qualifying as an overall oppressive system impeding attainment of the core values of development, menstruation stands out to drive the attention of policy makers.

Following the research on linkage between lack of menstruation hygiene and general women health, dialogues on menstruation has opened up at the policy level, introducing Menstrual Hygiene Scheme (MHS) in 2011 in selected 152 districts in India [10]. This paper, using unit level data from NFHS 4, identifies various sources of differential uptake of improved sanitary products in India, depending upon the duration of exposure of the girls in the program (based on their age), geographical location of the household (rural or urban) and the districts where they lived in. Using triple difference-in-difference [11,12] mechanism, the study finds that there is significant impact of shifting to hygienic products *only* among women with poorer education and low media exposure. The program could not mark any difference on behavioral changes in already better-off cohorts.

## Literature Survey

A predominantly large set of literature advocate for an urgent bridging of the existing knowledge gap in the arena of menstrual health as a solution [13–15]. The Intervention, Education, Communication (IEC) to follow, may take different forms of individualistic or community based approach [1,16]. Studies have compared the present scenario of Menstrual Hygiene Management (MHM) across countries and found that not many girls across developing countries knew about the menstruation process before menarche [14]. In primary surveys conducted in the respective countries, only 35% of girls in Bangladesh had reported a priori knowledge about the menstruation process and in India the corresponding share was 75.60%. However, untimely or inadequate sources of information are reported to be the mothers, sisters or friends. In Nepal, school text books are the most commonly reported source, while 75% of them had no menstrual information source. Parents find it inappropriate to talk about menstruation which is related to child birth and sex, letting it be a topic least talked about.

The concern even more aggravates for women in the developing countries due to lack of material resources for managing menstruation, like inadequacy of WASH facilities along with unaffordability of hygienic absorbents. Lack of hygienic menstrual products were found to be crucial in determining menstrual choice in low income countries like Kenya and have witnessed the onset of menstruation in turning girls into objects of sex [9]. Sex is often bartered for hygienic menstrual products. Studies in similar country setups highlights the need to improve infrastructure along with WASH facilities for better management of menstruation [16–18].

Given this backdrop of menstrual health scenario in developing countries, Government of India in 2011 had launched Menstrual Hygiene Scheme (MHS) [10] that used supplied subsidized sanitary pads and also generated awareness to attain the goal of improving menstrual health among adolescent girls in the rural area. Though the program seems quite holistic in terms of the intervention channels chosen, several studies have suggested that a single product solution without consideration of the context has often failed. While menstrual cup was a much celebrated solution for responsible period hygiene, an experiment in Uganda showed that the uptake of menstrual cup though free, wasn’t high. The reason being the lack of WASH infrastructure [4, 20]. Similar is the contrast of preference between re-usable pads and single-use pads. Where disposal is an issue, girls preferred reusable pads over single use products [21, 22] but in places that lacked adequate WASH facilities single use pads were preferred over reusable [23]. Solution to hygienic management of menstruation thus stands out to be contextual and dependent on the socio-cultural conditioning [4,21,24,25,26]. The paper so tries to examine if MHS has been the right mix of strategies opted to alter menstrual behavior and has located heterogeneity in the impact of the MHS for different cohorts.

## Materials and Method

### Data

The analysis uses secondary data from multiple sources. Data on menstrual choice, individual and other household characteristics was obtained from women’s questionnaire file of the 4^th^ round of National Family Health Survey, India (NFHS 4) carried out in the year 2015-16. Other district level controls like number of health centers was sourced from Rural Health Statistics in India, 2010 and nightlight data from the link http://blog.isharadata.com/2017/09/rnightlights-satellite-nightlight-data.html, respectively.

### Variables

The analysis uses a host of individual characteristics and district level characteristics to control for potential co-founding factors that affect menstrual health and to improve precision of estimates. The individual characteristics used as control are such as water source, toilet facility, religion, media exposure, beneficiary status under RSBY, status of health worker meet in last 3months, hemoglobin level, education and wealth index. The district levels control used are night light average and PHC per thousand population. Listed below are the variables that are redefined for the study purpose.

- *Media exposure* is categorized into two categories namely low exposure and high exposure. A low media exposed individual is defined as one who is not at all exposed to any one of the following: reading newspaper or magazine, watching television or listening to radio. Else an individual is assumed to be high media exposed.
- *Hemoglobin level* is also redefined to form a bivariate. An individual is marked as below normal if hemoglobin level is less than 12g/dl, above normal if otherwise.
- *Type of toilet facility* as defined in NFHS4 had multiple categories. Redefinition has now, identified toilet type into three broad categories: flush toilet, pit toilet and no facility/dry toilet, renaming the variable as toilet facility. Flush toilet category comprises of flush to piped sewer system, flush to septic tank, flush to pit latrine, flush to somewhere else, flush, don’t know where. Pit toilet category comprises of ventilated improved pit latrine (v.i.p.), pit latrine with slab and composting toilet. No facility/bush/field, dry toilet, other and not a de jure resident are compiled to form the third category.
- *Night light* average data collected over May 2012 to April 2013 is averaged and normalized over area per square kilometer.
- *Wealth index* is a composite measure of a household cumulative living standard. Uses household ownership of selected assets such as television, bicycles, materials used for housing construction, type of water access and sanitation facility.

Menstrual product usage which is the focal variable of study is at times classified into two types based on hygiene. The first category of hygienic menstrual product usage indicates usage of either locally made sanitary napkins or sanitary napkins or tampons and the second category of unhygienic menstrual product indicates usage of nothing/other or clothes for managing menses. Individuals using a mix of menstrual products are not included in the study. Only usage of exclusive menstrual product is used for the analysis.

### The program

Government of India in 2011 had launched Menstrual Hygiene Scheme (MHS) in rural areas of 152 selected districts and 21 states [10]. The scheme adopts two key strategies to improve menstrual health among girls of the age group 10-19years:

●Demand generation through ASHA and other community mechanisms such as Women’s Groups / *Kishori Mandals.* An additional mechanism for in-school youth would be that of the Adolescent Education Programme through the life skills courses for Classes IX and XI. (Kishori Mandal meetings are held to provide a platform for girls aged 11-16 to build their self-confidence by giving them inputs in life skills and information on topics they would not readily receive at home or in their school curriculum)
●Supply side intervention through ensuring a supply of a product (locally made sanitary napkin) which is reasonably priced and of high quality.

State criteria for selection of districts where this intervention was taken up were related to existing adolescent health programme, strong presence of Adolescent Education Programme(AEP) intervention, active Self-Help Group (SHG) federations, effective Accredited Social Health Activist (ASHA) training and support systems.

**Table 1** check for adherence to the district selection strategy using proxies indicating the spread of other government health programs. The proxies included meeting with Angawadi worker, ASHA or other health worker in the last 3 months, uptake of Rashtriya Swastha Bima Yojana (RSBY) and hemoglobin level. Hemoglobin level was used to capture the effect of “*The National Iron Plus initiative for Anemia Control among six months onward population*”. **Table 1(Panel A)** shows significant better utilization of health programs in the treated district, thereby confirming that the districts were well targeted as laid in the policy documents. The paper does not look into how equitable the MHS design was, but explores the effect of the scheme given the selection bias present in choosing districts for program implementation. Individual, household and district level characteristics (**Table 1: Panel A and B**) too show significant differences across the treatment status of districts. According to many indicators the treatment districts appeared to have better development indicators.

**Table 1.**
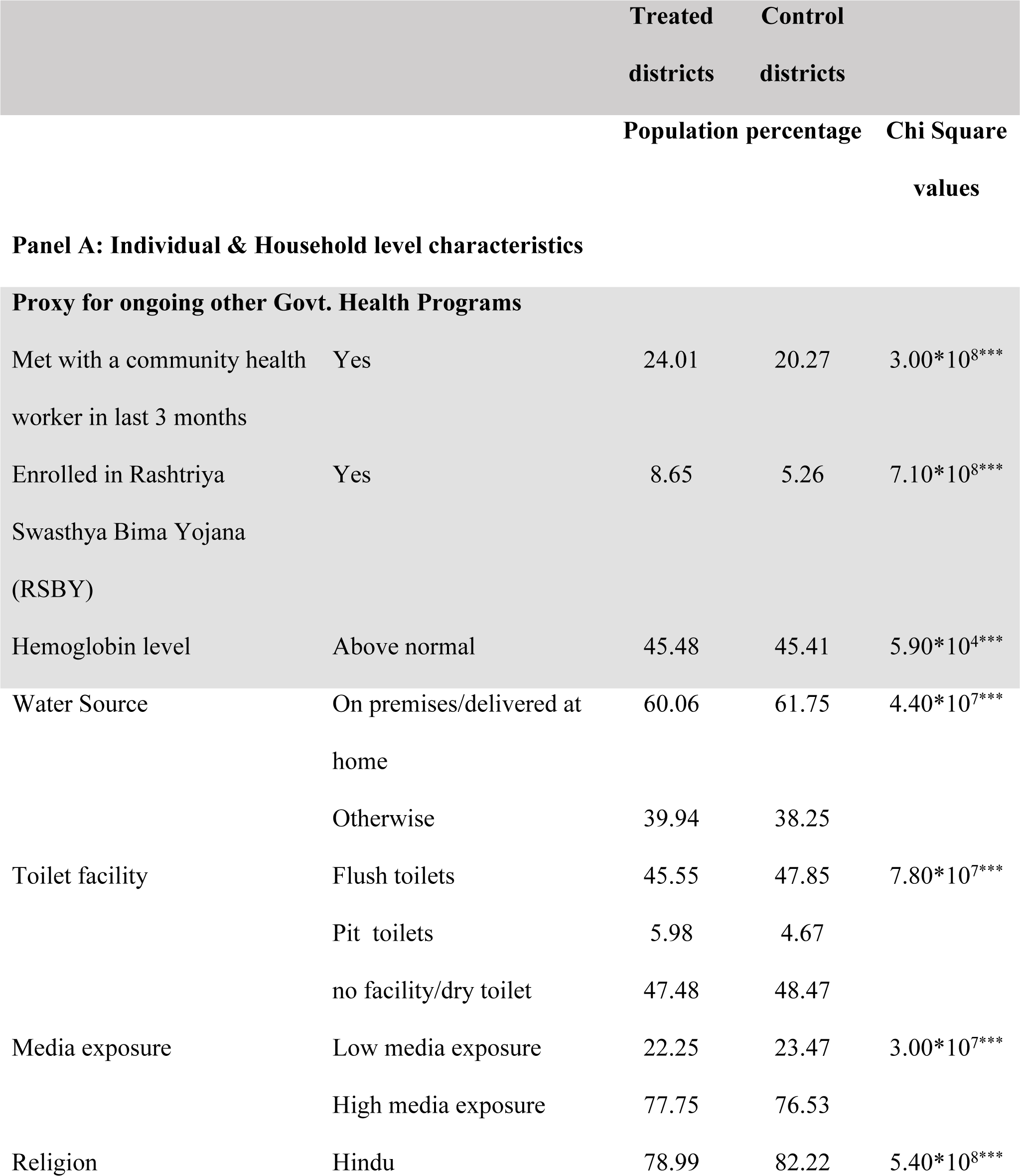
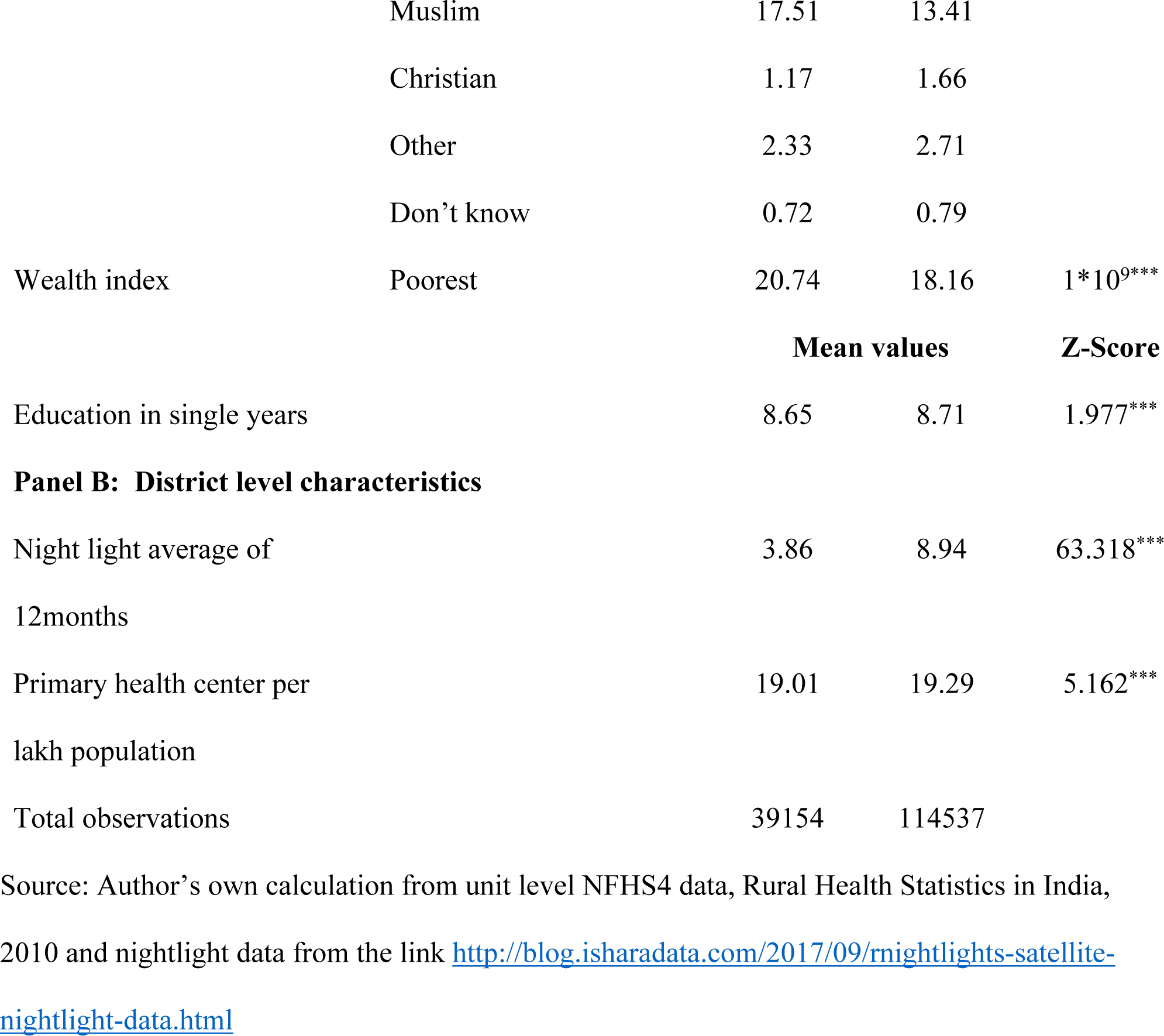
Composition of individual, household and district level characteristics across treated and control districts.

|  |  | **Treated**<br>**districts** | **Control**<br>**districts** |  |
| --- | --- | --- | --- | --- |
|  |  | **Population percentage** |  | **Chi Square**<br>**values** |
| **Panel A: Individual & Household level characteristics** |  |  |  |  |
| **Proxy for ongoing other Govt. Health Programs** |  |  |  |  |
| Met with a community health worker in last 3 months | Yes | 24.01 | 20.27 | $3.00*10^{8***}$ |
| Enrolled in Rashtriya Swasthya Bima Yojana (RSBY) | Yes | 8.65 | 5.26 | $7.10*10^{8***}$ |
| Hemoglobin level | Above normal | 45.48 | 45.41 | $5.90*10^{4***}$ |
| Water Source | On premises/delivered at home | 60.06 | 61.75 | $4.40*10^{7***}$ |
|  | Otherwise | 39.94 | 38.25 |  |
| Toilet facility | Flush toilets | 45.55 | 47.85 | $7.80*10^{7***}$ |
|  | Pit toilets | 5.98 | 4.67 |  |
|  | no facility/dry toilet | 47.48 | 48.47 |  |
| Media exposure | Low media exposure | 22.25 | 23.47 | $3.00*10^{7***}$ |
|  | High media exposure | 77.75 | 76.53 |  |
| Religion | Hindu | 78.99 | 82.22 | $5.40*10^{8***}$ |
|  | Muslim | 17.51 | 13.41 |  |
|  | Christian | 1.17 | 1.66 |  |
|  | Other | 2.33 | 2.71 |  |
|  | Don't know | 0.72 | 0.79 |  |
| Wealth index | Poorest | 20.74 | 18.16 | 1*10<sup>9</sup>*** |

|  |  | Mean values |  | Z-Score |
| --- | --- | --- | --- | --- |
| Education in single years |  | 8.65 | 8.71 | 1.977*** |
| **Panel B: District level characteristics** |  |  |  |  |
| Night light average of<br>12months |  | 3.86 | 8.94 | 63.318*** |
| Primary health center per<br>lakh population |  | 19.01 | 19.29 | 5.162*** |
| Total observations |  | 39154 | 114537 |  |

Source: Author's own calculation from unit level NFHS4 data, Rural Health Statistics in India, 2010 and nightlight data from the link [http://blog.isharadata.com/2017/09/nightlights-satellite-](http://blog.isharadata.com/2017/09/nightlights-satellite-nightlight-data.html) [nightlight-data.html](http://blog.isharadata.com/2017/09/nightlights-satellite-nightlight-data.html)

### Identification Strategy

The data used in the paper is from NFHS4 conducted in 2015-16, i.e. 5years after the roll out of MHS. Information for menstrual product usage is only available for girls between the age of 15-24yrs, which corresponded to MHS beneficiary cohort at the program initiation year. As the girls grew above 19years, they moved out of the program net, so the cohort 20-24years is no more currently exposed to the program. The program effect may seem to be lying in the difference of usage in hygienic menstrual product usage between the currently exposed 15-19years girls in the treated vis-a-vis the control district. Doing this would however over-report the effect of MHS by increased usage due to subsidized availability of sanitary napkins (supply effect). The effect thus doesn’t lie in assessing usage for currently exposed cohorts across treatment, but rather over behavioral change. Behavioral change takes time and is expected to work better with more time to treatment. We therefore use difference in duration of exposure for impact assessment. On being calculated since the roll-out of program in 2011, the 15-19years old cohort was exposed to the program for a full term of 5years and for the 20-24yrs cohort, exposure ranged in between 1- 5years. The 15-19years so classified into high exposed cohort and the 20-24years as the low exposed cohort.

The objective is to see if greater exposure to MHS has increased the probability of using hygienic menstrual product in the treated districts. The first difference compares this outcome between the high exposed cohort and the low exposed cohort of rural areas. However, this difference may be due to more instances of going out of the house during menstruation, found among the younger cohort on account of attending school or remaining within the program scope. We so use the urban cohort which was out of the program scope, as counterfactual to find second difference.

A systematic gap among the urban cohorts over usage is expected to be low even in the absence of MHS due to overall better awareness, compared to the rural cohorts. We therefore, construct a triple difference estimate of program impact by comparing the second differences of the treated district with the same second differences of control district. We have controlled for socio-economic variables and availability of healthcare facilities that are likely to affect menstrual choice. Hence, second differences for the districts can be expected to be same in absence of the program. Any difference in second differences (triple difference) can hence be inferred as the impact of MHS. The equation below models the triple difference through the coefficient, β_7_.

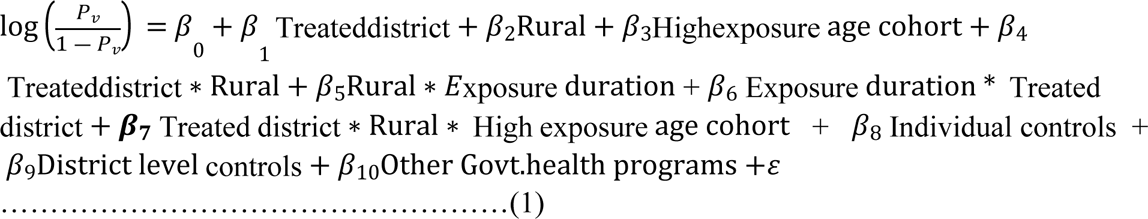

*P_V_* is the probability of using a particular variant (V^th^) of menstrual product, 1 being usage and 0 being non-usage (ordered logit).

## Results

### Menstrual product usage scenario of women in NFHS4

The choice of menstrual product for different age groups of women from NFHS 4 is shown in **Table 2**. Menstrual product usage differs across age cohorts and usage of hygienic menstrual products is found to be statistically higher (Chi-square= 3*10^6^) among the younger cohort. Cloth is the most popular choice of menstrual product with 53.89% usage, followed by a 33.27% usage of sanitary napkins. Nothing and others are the least opted choice for menstrual management, whereas, locally made sanitary napkins and tampons are used moderately by women in the sample. Unhygienic menstrual product which comprises of cloth and nothing/other is used by more than half of the population while the use of hygienic menstrual product i.e. locally made sanitary napkins, sanitary napkins and tampons is used by only 45.43% of the population. The usage of hygienic products is more among currently exposed cohort of 15-19 years, rather than those who have shifted out of the program net.

**Table 2.** Choice of menstrual product among women in NFHS4(%).

| Choice of menstrual product | 15-19 Years | 20-24 Years | 15-24 combined |
| --- | --- | --- | --- |
| Nothing /Other | 0.64 | 0.72 | **0.68** |
| Cloth | 53.72 | 54.05 | **53.89** |
| Locally made sanitary | 11.02 | 10.62 | **10.82** |
| Sanitary napkins | 33.29 | 33.26 | **33.27** |
| Tampons | 1.32 | 1.35 | **1.34** |
| **Total** | **100** | **100** | **100** |
| **Hygienic product** | 45.64 | 45.23 | **45.43** |
| **Unhygienic product** | 54.36 | 54.77 | **54.57** |
| **Total sample size** | 77275 | 76416 | 153,691 |

Source: Author's own calculation from unit level NFHS4 data

### Exploring the difference in menstrual product usage across individual and household categories

A look through into **Table 3** identifies that apart from age, other individual, household and district level characteristics of women also serves to be a basis of differential menstrual hygiene management choice. Differences in choice of menstrual product across variable categories is evident. High media exposure, high level of education, improved flush toilet facility and supply of water on premises are factors that positively affect usage of hygienic menstrual products. These findings reiterate the importance of bridging the knowledge gap and improving WASH facilities to ameliorate menstrual hygiene management decision.

**Table 3.**
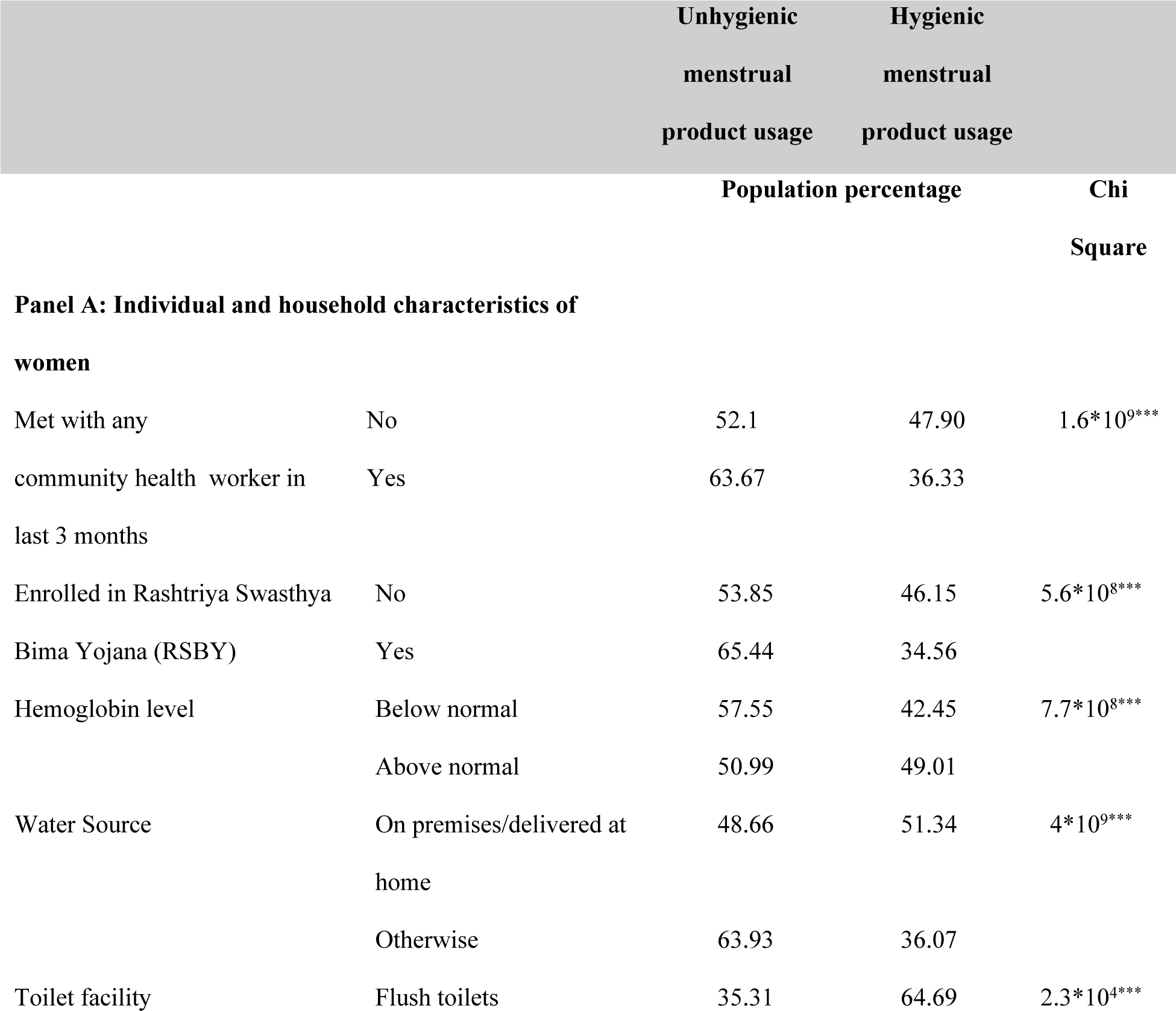
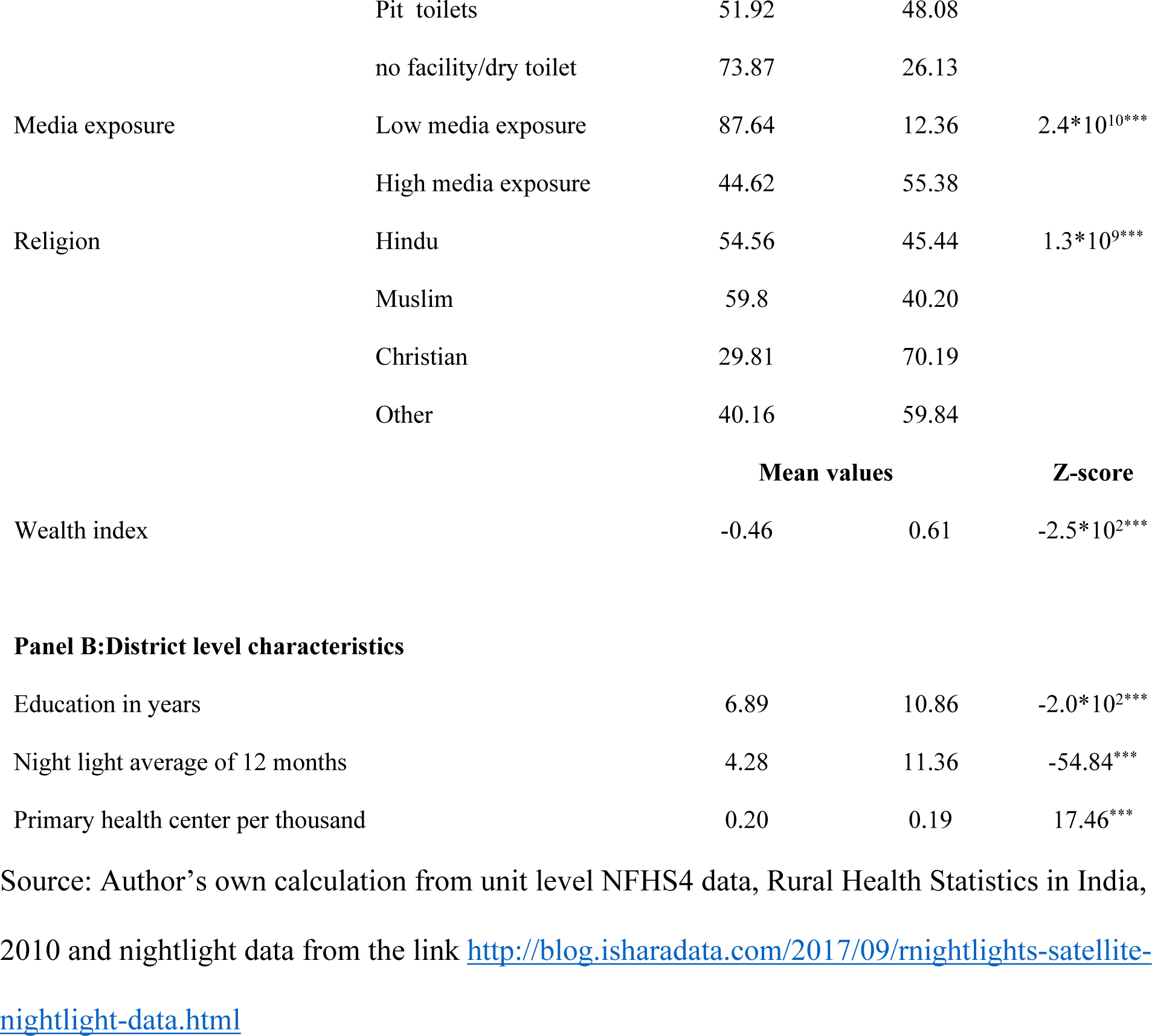
Difference in menstrual product usage within individual, household and district level variables.

We have also explored the difference in usage for socio-demographic variables such as religion, owing to close association of social taboos and local knowledge with menstrual practices. Plotting differences in such practices over religion reveals that a higher share of Christians uses more hygienic menstrual product in comparison to the rest of the religions. Additionally, places with high economic activities and richer individuals are seen to be using more hygienic menstrual products.

Interesting results surface across differences in health care utilization. Women who has reported greater interaction with health workers and enrollment under RSBY scheme, are the ones with higher usage of unhygienic menstrual products. Being over-deterministic of various other individual and household characteristics, the simple descriptive statistics results (and not correlations) do not infer on the direction of usage of menstrual products.

The sample characteristics of individuals stylize the treated districts by higher proportion of factors that contribute towards a more unhygienic management of menstruation such as use of pit toilets, water source outside premises, higher share of Muslims, lower level of education, lower economic activities, higher share of women meeting health workers in the last 3 months and higher exposure to RSBY in treated districts, which is a indication to targeting districts that performed poorer in respect to menstrual hygiene. Our exercise of triple difference so controls for these individual, household and district level characteristics when comparing MHS impact between the treated and control districts.

### Triple difference-in-difference results

Triple difference in difference measures the MHS impact by looking at the menstrual choices of individual who has been exposed to the program for a longer time and belongs to rural area of the treated district using the β_7_coefficient of the estimated equation. The estimation equation has been used with individual and district level health facility controls that otherwise would show a biased menstrual choice results for the selected districts. Descriptive statistics find that a well laid out district selection strategy for program roll-out in the policy document has carefully encompassed those, where adolescent program was already on the go and created higher chances of already hygienic menstrual product usage even in absence of MHS intervention, through channels of health awareness and thus calls for controls.

The MHS impact as captured through D3 in **Table 4** clearly indicates that no significant alteration in the product choice was found over MHS coverage. Usage of all kind of menstrual products remained unchanged for the treated group with longer exposure. The sample as a whole finds MHS to be ineffective in bringing about any significant behavioral change.

**Table 4.**
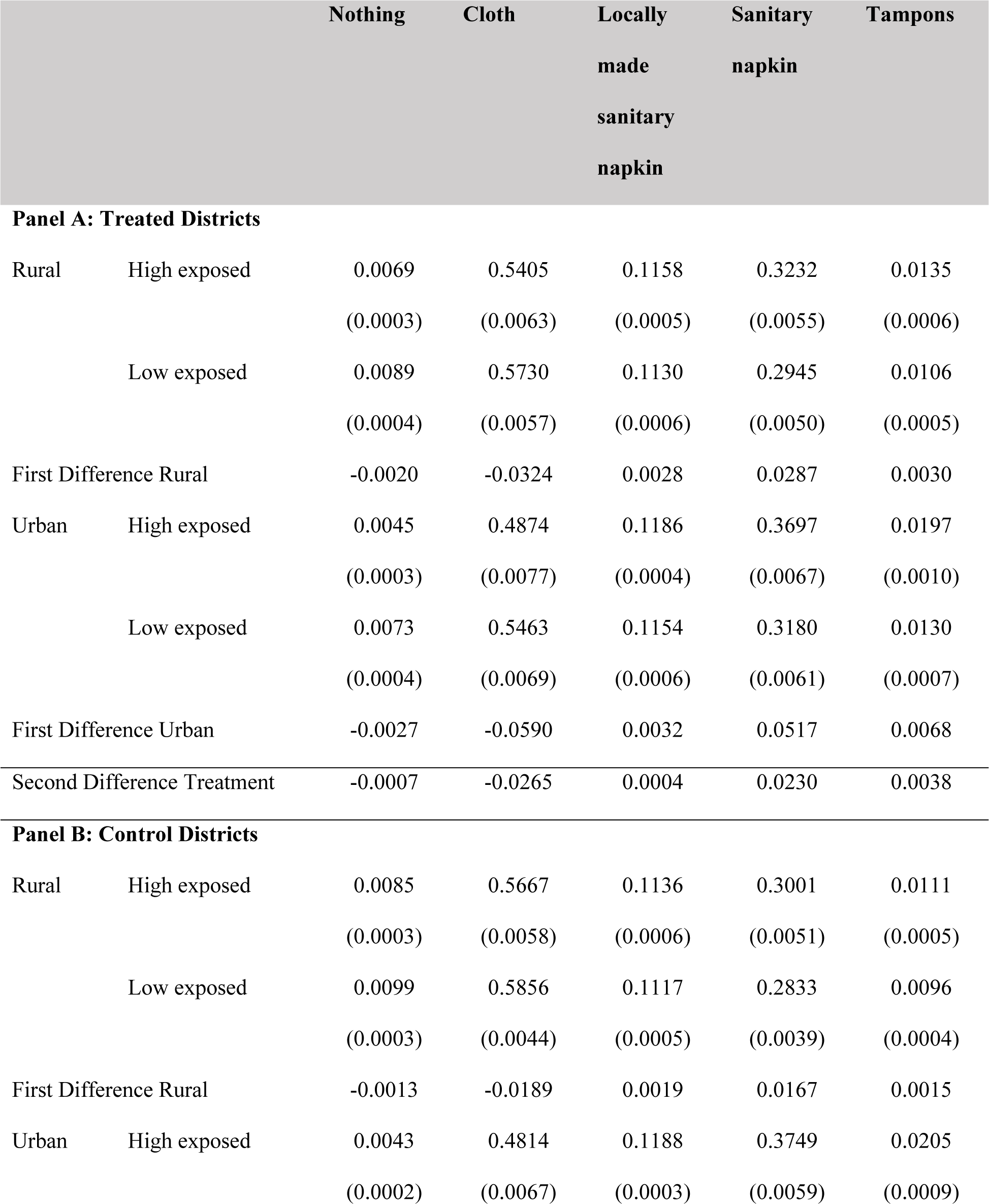
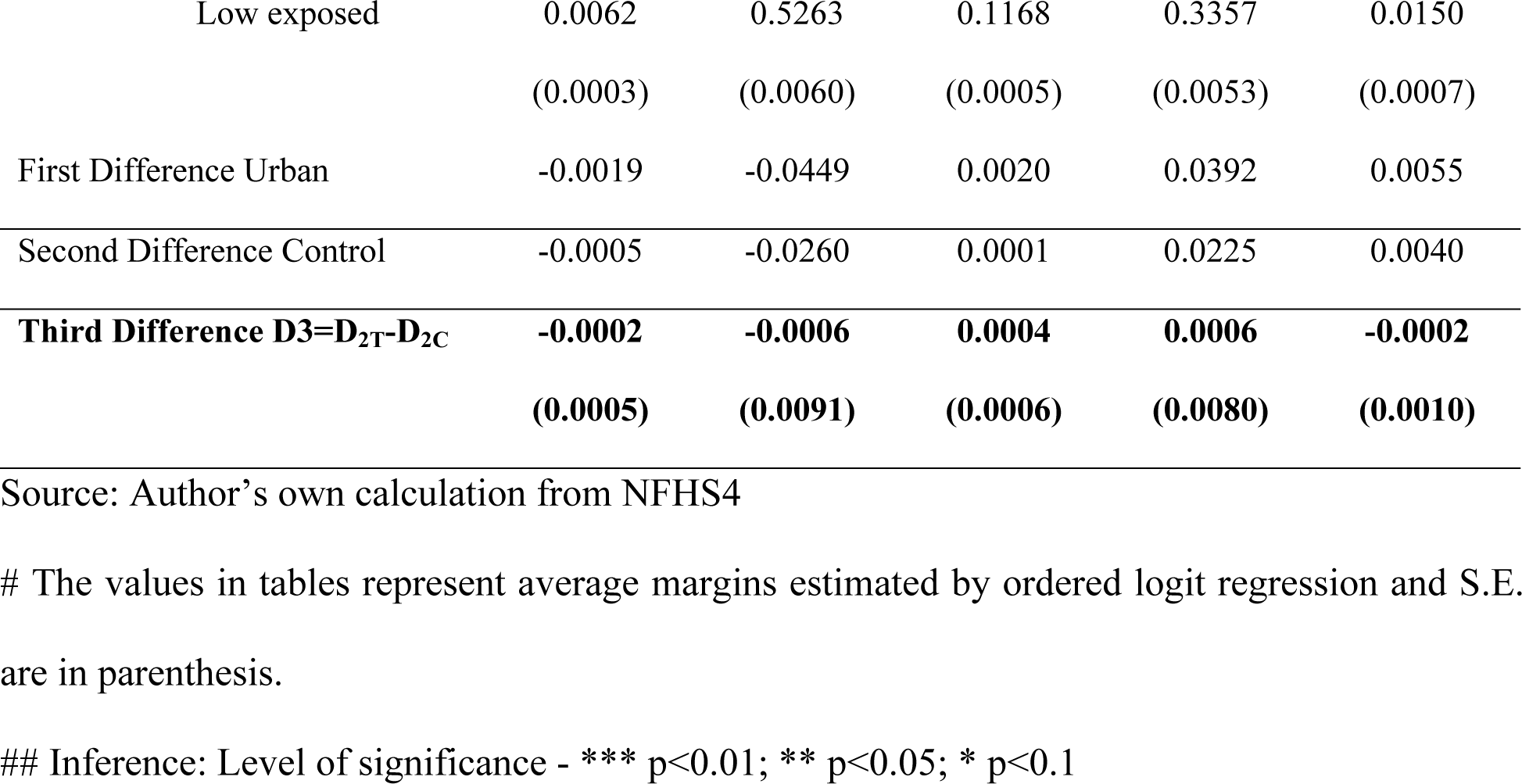
Triple difference results showing impact of MHS on usage of different menstrual products.

### Heterogeneity check for MHS effect

MHS might however be found to have differential effect over cohorts based on their socio- economic characteristics. As already found in literature, contextual parameters play an important role in determining one’s adaptability to a product. We explore the MHS effect through triple difference-in-difference as earlier but for different cohorts. The choice of socio-economic variables was to see if the interventions were well targeted towards unaffordability and lack of awareness, we so divided the cohort according to their wealth, media exposure and education (**Table 5**). We find that individuals with no exposure to mass media saw all kind of shifts to hygienic menstrual product as an impact of MHS. Effect of MHS was absent among the girls with high media exposure. Only changes in usage of locally made sanitary napkins was observed, standing at a low of 0.09% increased probability. Moving on to the next variable used as proxy for awareness, education was dichotomized into below 10years education and above 10 years of education. Triple difference in difference results show that there was a significant shift to sanitary napkins (both locally made and others) from cloth and no protection for the individuals with less than 10years of education, only. The cohort with higher education saw no significant changes in usage of any of the menstrual product.

**Table 5:** Cohort-wise triple difference (D3) results showing impact of MHS on usage of different menstrual products.

|  | PANEL A |  |  | PANEL B |  |  |
| --- | --- | --- | --- | --- | --- | --- |
|  | No exposure to mass media |  |  | Exposed to mass media |  |  |
|  | D<sub>3</sub> | Std. Err. | P>chi2 | D<sub>3</sub> | Std. Err. | P>chi2 |
| Nothing | -0.0062*** | 0.0023 | 0.0077 | 0.0001 | 0.0003 | 0.8335 |
| Cloth | -0.0642*** | 0.0223 | 0.0040 | 0.0076 | 0.0113 | 0.5038 |
| Locally made sanitary napkins | 0.0167*** | 0.0057 | 0.0034 | 0.0009*** | 0.0004 | 0.0161 |
| Sanitary Napkins | 0.0490*** | 0.0170 | 0.0039 | -0.0076 | 0.0106 | 0.4751 |
| Tampons | 0.0047*** | 0.0017 | 0.0061 | -0.0009 | 0.0009 | 0.2846 |
|  | Education less than 10yrs |  |  | Education more than 10yrs |  |  |
|  | D<sub>3</sub> | Std. Err. | P>chi2 | D<sub>3</sub> | Std. Err. | P>chi2 |
| Nothing | -0.0025*** | 0.0009 | 0.0081 | 0.0006 | 0.0004 | 0.1712 |
| Cloth | -0.0234* | 0.0130 | 0.0719 | 0.0141 | 0.0127 | 0.2693 |
| Locally made sanitary napkins | 0.0055*** | 0.0025 | 0.0244 | 0.0010 | 0.0016 | 0.5382 |
| Sanitary Napkins | 0.0192* | 0.0105 | 0.0675 | -0.0141 | 0.0128 | 0.2697 |
| Tampons | 0.0012 | 0.0010 | 0.2184 | -0.0015 | 0.0019 | 0.4356 |
|  | Wealth below mean |  |  | Wealth above mean |  |  |

|  | D<sub>3</sub> | Std. Err. | P>chi2 | D<sub>3</sub> | Std. Err. | P>chi2 |
| --- | --- | --- | --- | --- | --- | --- |
| Nothing | 0.0015 | 0.0011 | 0.1804 | -0.0005 | 0.0005 | 0.3685 |
| Cloth | 0.0362* | 0.0191 | 0.0577 | -0.0186 | 0.0125 | 0.1356 |
| Locally made sanitary napkins | -0.0065* | 0.0038 | 0.0874 | -0.0033* | 0.0017 | 0.0526 |
| Sanitary Napkins | -0.0277* | 0.0147 | 0.0593 | 0.0194 | 0.0127 | 0.1276 |
| Tampons | -0.0035** | 0.0017 | 0.0400 | 0.0030 | 0.0018 | 0.1026 |

Source: Author's own calculation from NFHS4 using ordered logit
#D<sub>3</sub> represents average margins estimated using ordered logit regression.

Dichotomizing the wealth variable gives us interesting results. It is seen that usage of sanitary napkins post-MHS intervention for the targeted has decreased while increasing the usage of cloth by 3.6% for the less wealthy individuals. Even though similar pattern of decreased hygienic product usage was observed for the richer individuals, it was almost insignificant in magnitude. The usage of locally made sanitary napkins decreased by 0.3% for the richer cohort.

## Conclusion

The paper finds that controlling for most of the characteristics that guide menstrual hygiene to minimize the self-selection into program bias, renders MHS ineffective. The program has failed to bring significant behavioral change among adolescent girls. With a limited time, product support the cohort seems to resort back to their initial choice and behaves much like the non-treated cohort. Even longer exposure to the program could not alter the menstrual behavior of individuals. The high reported percentage of using hygienic menstrual product in NFHS4 can so be seen as not an effect of Government interventions but an increase in overall development trend. Development of an economy is stylized by certain events like market expansion in remote areas and increased exposure to information through greater access to mobile phones which are also conducive to increased hygienic menstrual usage. Over the years, these things might have improved the product choice with and without MHS. However, saying this would be too simple to consider MHS as completely ineffective. A careful dive into the design of MHS strategies would help us understand that affordability and awareness was targeted to improve menstrual hygienic condition. The current analysis so used proxies for this target characteristics and divided the cohort accordingly. It is seen that the individuals with low education (less than 10years) and low media exposure has experienced the behavioral change owing to MHS, indicating that though the development activities were conducive to increased usage, not all could reap benefits of the same. A strategy like MHS addressed the marginalization of the cohort with lower awareness and altered their menstrual behavior. The maximum shift being to sanitary napkins.

**Table 5** shows that with longer exposure to MHS, the cohort with wealth below the mean has shifted to using cloth from sanitary napkins and tampons, whereas the counterpart shows no such impact. The impact of MHS thus seems to have adversely affected the poorer cohort and appears confusing. “*Period poverty*” has been the most highlighted problem associated with menstrual hygiene in developing countries like India and MHS had rightly tried to address the issue by providing subsidized napkins. The effect may however seem to vanish as the cohorts move out of the benefit net. Any behavioral impact of MHS on the cohorts with different wealth levels so shouldn’t be assessed in the light of increased usage of marketed goods. The reverse results might so be interpreted as a shift to using traditional methods of protection like cloth in a hygienic manner, which would rather be sustainable even in the absence of program. We might so conclude that MHS has rightly filled in the knowledge gap and helped girls to resort to hygienic and sustainable changes.

## Policy prescription

As MHS was found to be effective only for cohorts with lowest awareness, a proper targeting of cohorts should be done to considerably reduce the program cost and free the scheme of supply side fund constraints. As the marginalized section actually needed the program to shift to hygienic choices, more concerted efforts should be targeted towards them. Further, the fund saved could be reallocated to include older cohorts under the program scope, so that they do not slip back to unhygienic products just because of poverty. Also with a reduced sample of beneficiaries IEC interventions are likely to be implemented with greater effectiveness and contribute in increasing the probability of using hygienic products by a greater extent.

## Data Availability

If the data are all contained within the manuscript and/or Supporting Information files, enter the following: All relevant data are within the manuscript and its Supporting Information files.

## Acknowledgement

The authors thankfully acknowledge the insightful advices from Ishita Mukhopadhyay (Professor, Department of Economics, University of Calcutta) and S Anukriti (Senior Economist, Development Research Group, The World Bank, Washington DC) as members of Research Advisory Committee of author, Ms. Olivia Sarkar.

## Ethical Statement

The study is based on freely downloadable secondary NFHS4 data, obtained from Demographic Health Survey (https://dhsprogram.com/methodology/survey/survey-display-355.cfm) and hence doesn’t have ethical compliances.

## Notes

### Competing Interest Statement

The authors have declared no competing interest.

### Funding Statement

The author(s) received no specific funding for this work.

